# A protocol for a systematic review and individual patient data meta-analysis investigating the relationship between *Pfkelch13* mutations and response to artemisinin-based treatment for uncomplicated falciparum malaria

**DOI:** 10.1101/2025.02.05.25321713

**Authors:** Stephanie van Wyk, Prabin Dahal, J. Christevy Vouvoungui, Dhol S. Ayuen, Farhad Shokraneh, Soma Aboubakar, James A. Watson, Phillipe J Guerin, Karen I Barnes

**Author notes:** Correspondence: Prof Karen Barnes.

## Abstract

**Introduction:** Artemisinin-based combination therapies (ACTs) remain the WHO-recommended treatment for uncomplicated *Plasmodium falciparum* malaria. However, the emergence and spread of artemisinin resistance (ART-R) threatens ACT efficacy. ART-R is phenotypically expressed as delayed parasite clearance, which can facilitate ACT partner drug resistance. ART-R has been causally linked to specific mutations in the *Pfkelch13* gene.

**Methods and Analysis:** The systematic review and associated meta-analysis aim to determine the correlation between *Pfkelch13* genotypes and clinical and parasitological response to ACTs from a globally representative dataset pooling individual patient data (IPD) from eligible published and unpublished studies. The eligibility criteria include *Pfkelch13* genotyping results at baseline complemented by individually linked parasitological and clinical assessments following artemisinin-based treatment. The data will be curated, standardised, and analysed using this proposed statistical analysis plan, adhering to PRISMA-IPD guidelines. Our statistical analysis plan (SAP) will apply hierarchical modelling to assess the effect of *Pfkelch13* mutations on parasite clearance half-life and therapeutic efficacy across different regions. This will include study sites as random effects in the model and potential predictors such as age, sex, baseline parasite load and other potential effect modifiers as fixed effects. This analysis will enhance the understanding of the influence of *Pfkelch13* mutations on malaria treatment outcomes.

**Ethics and Dissemination:** Data is obtained with informed consent and ethical approvals from the relevant countries and was pseudonymised before curation in the IDDO/WWARN repository. Data ownership remains with contributors. This IPD meta-analysis met the Oxford Tropical Research Ethics Committee criteria for waiving ethical review, as it is a secondary analysis of existing pseudonymised data. The resulting peer-reviewed publication will help strengthen and enhance the efficiency of ART-R surveillance and response and support policy decisions. The results will be submitted for publication in a peer-reviewed journal and shared at scientific conferences.

**Registration details:** PROSPERO-registered (CRD42019133366)

**Article summary:** *Strengths and limitations of study:*

1. Inclusion of all available data based on a PROSPERO-registered systematic review of published and unpublished studies representing diverse patient and malaria parasite populations across varied global geographic regions, malaria transmission intensities and antimalarial drug pressures.
2. Individual patient data (IPD) meta-analysis is the only valid approach to determine the relationship between mutant alleles and parasite clearance, taking into account moderators of treatment response. This protocol outlines the combined efforts of stakeholders, national malaria programs, and researchers to investigate this relationship through data reuse.
3. This WWARN Data Management and Statistical Analysis Plan allows for standardised IPD to be included in the WWARN repository, which is then pooled into a single, quality-assured database using Clinical Data Interchange Standards Consortium (CDISC) standards.
4. IPD meta-analyses are limited by the data available, with inconsistencies in the type and completeness of covariates measured across the studies. For example, studies often only assess known mutations previously associated with ART-R, reducing the likelihood of detecting clinically relevant mutations not previously reported.
5. A key limitation is that current published studies may be insufficient to investigate the direct impact of *Pfkelch13* mutant genotypes on the predictors/indicators of increased malaria transmission.

## Introduction

Artemisinin-based combination therapies (ACTs) are the cornerstone for treating uncomplicated *Plasmodium falciparum* malaria globally. These therapies combine a fast-acting artemisinin derivative with a longer-acting partner drug [1–3]. In Southeast Asia, the efficacy of most first-line Artemisinin-based Combination Therapies (ACTs) has significantly declined due to widespread resistance to both the artemisinin component and its partner drugs. This resistance emerged initially against artemisinin, known as “partial” ACT resistance (ART-R), with resistance to the partner drugs following closely thereafter. The situation is becoming increasingly concerning in Africa, where the rapid emergence and spread of artemisinin resistance now threatens the effectiveness of ACTs. This is particularly alarming given that Africa accounts for the majority of global malaria cases and fatalities. If first-line ACTs fail in sub-Saharan Africa, particularly Artemether-Lumefantrine (AL), which constitutes 70% of all administered treatments on the continent, this would pose a significant challenge to global malaria control and elimination efforts [1, 2].

ART-R is partly driven by non-synonymous mutations in the *P. falciparum Kelch 13* (*Pfkelch13*) gene, while potential other mutations may have a synergistic effect [2, 4, 5]. These mutations result in the loss of ring-stage sensitivity of the parasites to artemisinin, resulting in delayed parasite clearance following the start of treatment. This is characterised by an increased parasite clearance half-life (PC½) [6, 7]. Consequently, fewer parasites are killed by the artemisinin derivative over the 3-day course (two parasite life cycles), thus increasing the burden on the partner drug to eliminate residual parasites. This undermines the efficacy of the 3-day ACT treatment [6].

The *Pfkelch13* non-synonymous mutations underlying clinical and experimental ART-R include the BTB/POZ and propeller domains (**Table 1**). Over 260 *Pfkelch13* non-synonymous mutations have been reported. However, only about 25 of these mutations are associated with the clinical phenotype of ART-R, *i.e*., prolonged parasite clearance. They are considered markers used for genetic surveillance to monitor the emergence of ART-R globally [4, 5, 8]. Concerning markers in Southeast Asia region include C580Y, R539T, and F446I, whereas P441L, C469F, C469Y, R561H, R622I, and A675V are prevalent in East Africa [5]. The World Health Organization (WHO) has classified genetic markers of concern into candidate and validated markers of ART-R [8, 9]. The *Pfkelch13* mutant markers that are associated with delayed parasite clearance or increased survival during ring-stage (observed as RSA^0–3h^) or differential effects in isogenic lines compared to wild-type are termed candidate markers of resistance, whereas validated markers of resistance have both experimental and clinical evidence of resistance [9]. This WHO classification system guides genomic surveillance initiatives to determine their geographical prevalence in malaria-endemic regions. Therefore, a comprehensive understanding of the genetic determinants driving ART-R will facilitate public health responses in these regions.

**Table 1:**
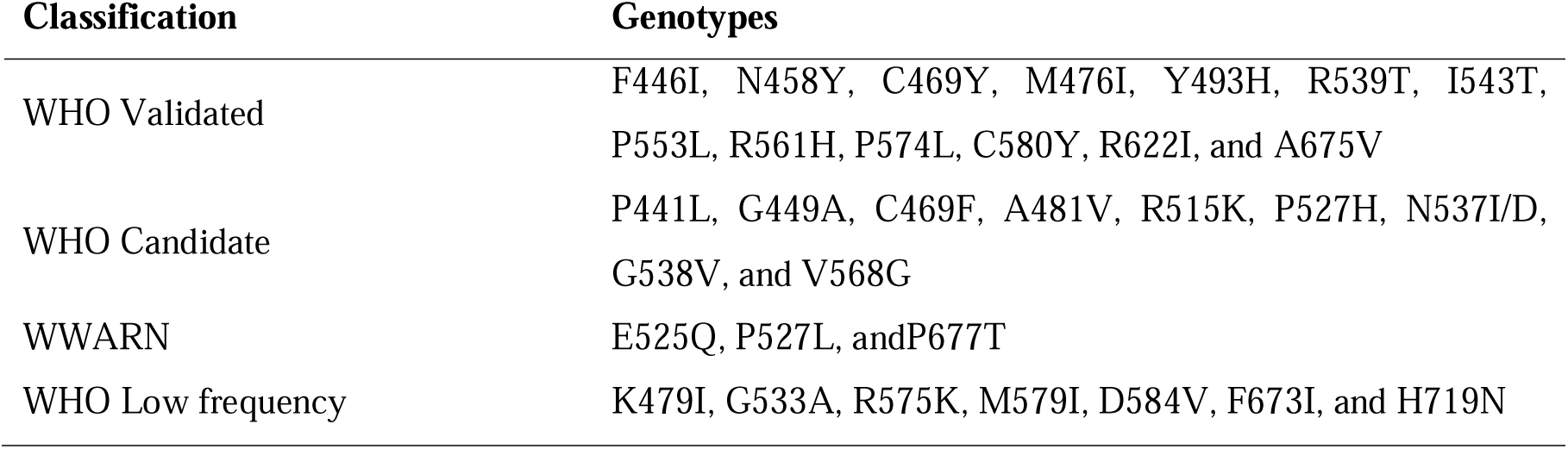
Summary of *Plasmodium falciparum* non-synonymous mutations of the *PfKelch13* gene regions and their associated classifications (1, 2).

Prior research has investigated the association between *Pfkelch13* markers and the rates of parasite clearance post-artemisinin treatments, identifying *Pfkelch13* markers significantly associated with slower parasite clearance in populations of the Southeast Asian region [8, 10] before the detection of ART-R in sub-Saharan Africa [11]. However, recent reports of *de novo* emergence and spread of ART-R in Africa [4, 5, 12], particularly in East Africa [13–21] and the Horn of Africa [22–27], are of enormous concern [3, 28, 29], warranting an updated investigation into the global prevalence, distribution, functional and clinical significance of *Pfkelch13* non-synonymous mutations [5, 15, 19, 28–33].

In sub-Saharan Africa, AL is predominantly utilised as the first-line ACT treatment for malaria and, for many countries, represents the only available treatment option [2]. However, concerns arise as therapeutic efficacy studies (TES) have demonstrated diminished efficacy of AL in Uganda, the Democratic Republic of the Congo (DRC), Kenya, and Tanzania [4, 36–39], falling below the WHO’s efficacy threshold (**Figure 1**). In these countries, the increased treatment failure often coincided with increased prevalence of ART-R. There are further concerns about the outdated nature of available TES data in many countries [2, 4, 9, 15, 36–40]. A pooled meta-analysis using robust statistical methods is imperative to bridge these information gaps and better understand the dynamics of *Pfkelch13* markers, the emergence of ART-R, and reduced ACT efficacy.

**Figure 1:**
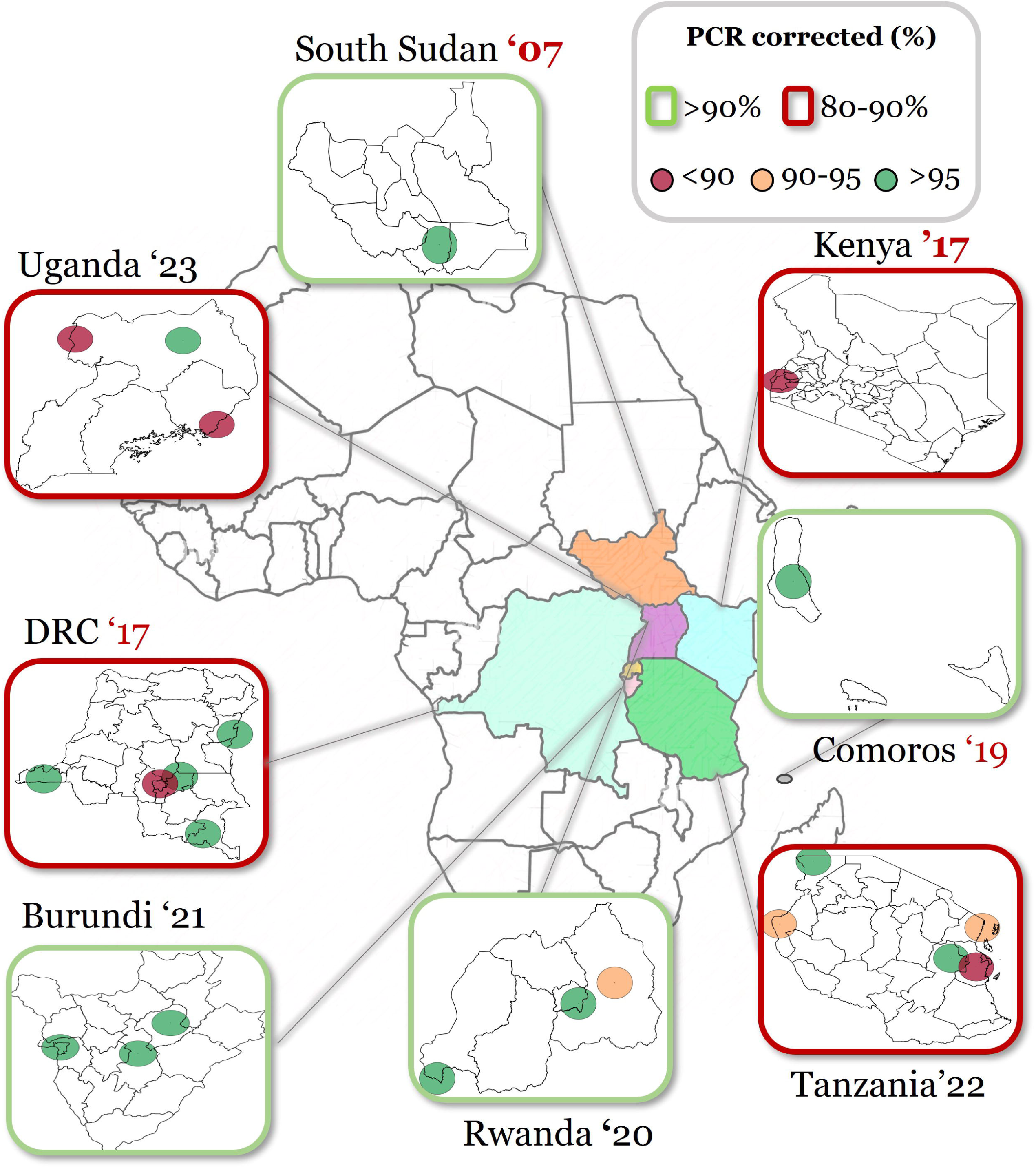
Outcomes of therapeutic efficacy studies of Artemether-lumefantrine in Eastern Africa. The figure indicates the PCR-corrected adequate clinical and parasitological response (ACPR) percentages per sentinel site. Countries outlined in red had one or more sentinel sites below the WHO threshold of 90% (80-90%); those above 90% are indicated in green. The colour-coded dots represent the PCR-corrected ACPR outcomes results and the sentinel sites’ geographic location per country. APCR rates below 90% are indicated in red, between 90-95% in orange, and above 95% in green, as indicated in the figure legend.

To address these evidence gaps, we propose this systematic review methodology and Statistical Analyses Plan (SAP), which will allow for undertaking individual participant data (IPD) meta-analysis (IPD-MA) by incorporating all relevant data collected since 2014. This protocol aims to generate a reproducible model to support subsequent analyses and continue to inform on the influence of *Pfkelch13* markers on the global ART-R landscape. This PROSPERO registered protocol adheres to PRISMA-IPD guidelines, utilising standardised methodologies and providing detailed definitions. The analysis aims to comprehensively evaluate the global prevalence, geographic distribution, and functional and clinical impacts of *Pfkelch13* markers on the outcomes of treatment for uncomplicated malaria. In turn, these findings will enhance the current understanding of the genetic determinants contributing to ART-R and its facilitation of reduced ACT efficacy and strengthen and enhance the efficiency of ART-R surveillance. The specific aims of this proposed IPD MA include:

1. To define the functional significance of *Pfkelch13* propeller non-synonymous mutations in patients with uncomplicated falciparum malaria treated with ACTs or monotherapy by location, treatment, study population and date in terms of:

□ Parasite clearance half-life (PC½).
□ Parasite positivity on Day-2 and Day-3.
□ Clinical and parasitological treatment response at the end of the study follow-up (Day-28, Day-42, or Day-63).
2. To compare the global prevalence and distribution of *Pfkelch13* mutations between different geographical regions, such as Asia and Africa, and intra-regional comparisons, such as Eastern and Southern Africa.

## Methods and Analysis

### Overview of Approach

This systematic review protocol details the methodologies for a comprehensive search and screening strategy and meta-analysis, leveraging an SAP for IPD to understand the association between *PfKelch13* markers and their phenotypic presentation (**Figure 2**). This proposed IPD-MA adheres to the PRISMA-IPD guidelines to integrate published and unpublished clinical data globally.

**Figure 2:**
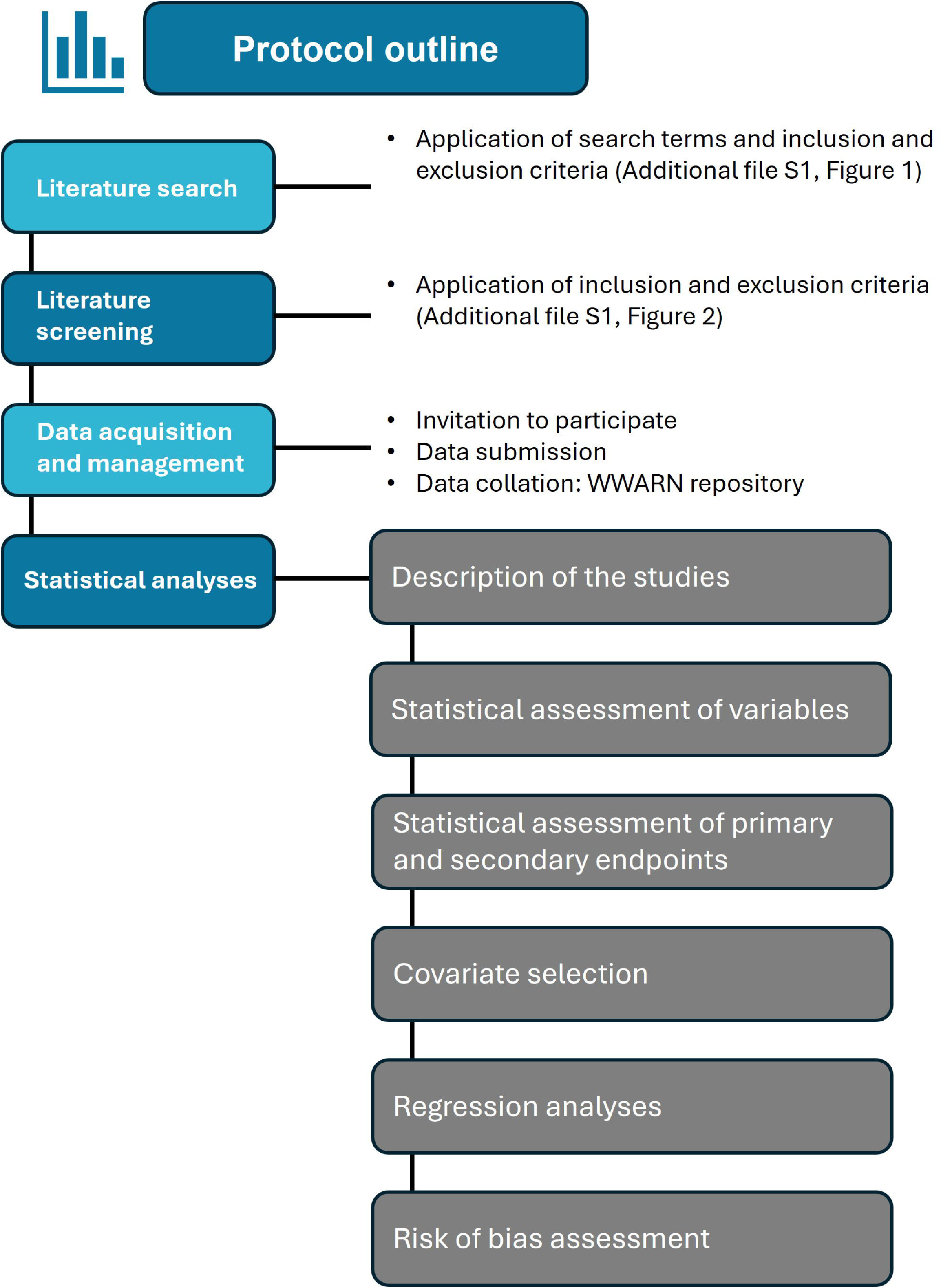
Outline of the protocol for investigating the relationship between *Pfkelch13* non-synonymous mutations and parasite clearance half-life in uncomplicated falciparum malaria through a systematic review and individual patient data meta-analysis.

### Literature search

The literature search strategy seeks published research articles and unpublished clinical trials from several bibliographical databases and clinical and research data repositories, detailed in **Supplementary Table S1** and outlined in **Figures 3** and **4**. Inclusion criteria are based on the availability of clinical patient characteristics and linked data, namely parasite clearance rates and treatment outcomes, and link genotypic data from the parasite responsible for the infection (**Table 2**). Exclusion criteria contain studies focusing solely on non-malarial, prophylactic, or herbal interventions (**Table 2**). Literature searches included publications and trials reported between 1^st^ January 2014 and 31^st^ August 2024. Search terms per repositories, databases, and libraries used are listed in **Supplementary material S2**, and further details can be found on PROSPERO (CRD42019133366) and associated publications Takata *et al.,* (2020) [34].

**Figure 3:**
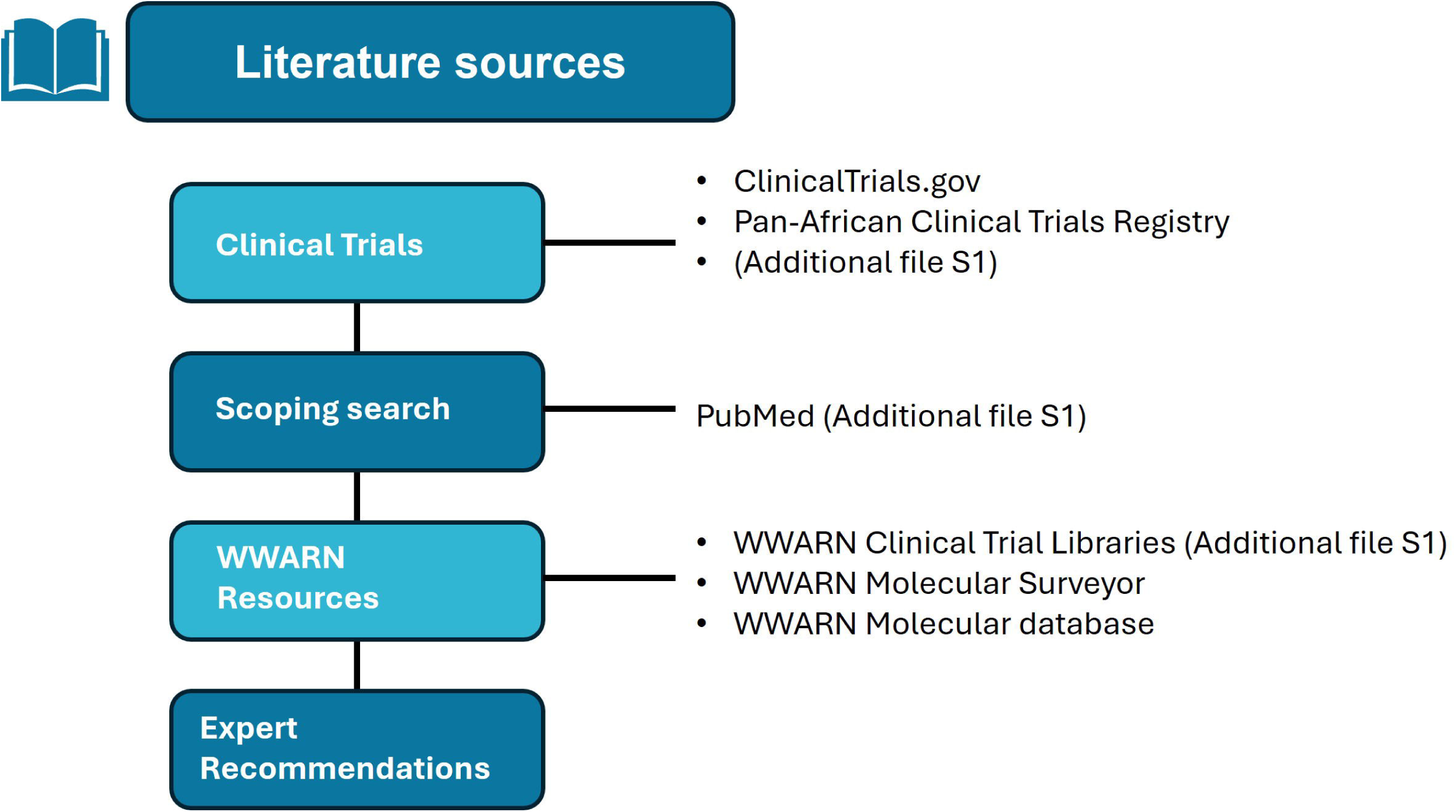
Outline of literature sources included in the literature search strategy.

**Figure 4:**
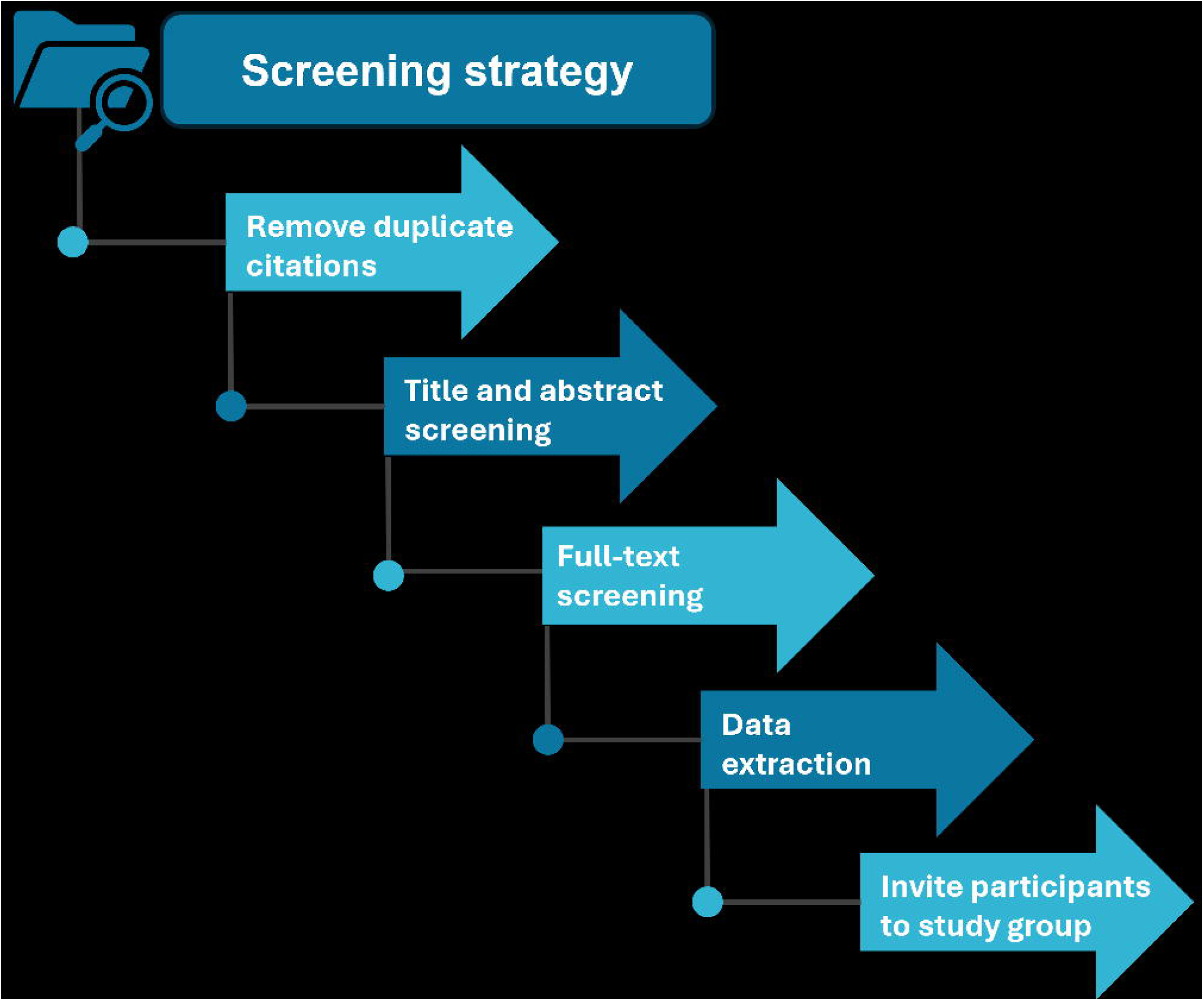
Outline of the screening strategy applied to determine eligible studies included in the review

**Table 2:** Summary of the literature search inclusion and exclusion criteria.

| Inclusion Criteria | Exclusion Criteria |
| --- | --- |
| • Studies assessing the efficacy of artemisinin-based antimalarials<br>• A minimum follow-up of two days<br>• Publications in any language<br>• Review articles | • Mass drug administration studies<br>• Herbal medicine studies<br>• Animal studies (non-human)<br>• Duplicate studies<br>• Case reports, case series, opinion pieces, and meta-analyses<br>• Retrospective studies<br>• Conference abstracts<br>• Pooled analyses or IPD meta-analyses specifically on *PfKelch13* and phenotype studies<br>• Studies focusing solely on safety<br>• Severe malaria-only studies |

#### Literature screening

**Figure 3** and **Supplementary table S1** outline the sources of literature used to identify relevant records. The screening process will involve removing duplicate references, screening studies by title and abstract, and reviewing studies in full text, and this will be followed by extracting data from records and inviting authors of the eligible studies to participate in the study group. Software used for the screening process includes Endnote (2023, version 20) and Covidence (2023; Covidence systematic review software). Endnote collates the studies identified through the literature search strategy, and references from Endnote are imported into Covidence. At least two independent reviewers will review the records identified during abstract, title, and full-text screening. Conflicts are resolved through a third independent reviewer.

The inclusion and exclusion criteria are outlined in **Table 2**. The following studies will be excluded: studies without information on drug regimens, treatment regimens not involving ACT or artemisinin derivatives, absence of data on *Pfkelch13* genotyping, or no serial parasite measurements undertaken. We also excluded review articles and studies that reported exclusively wild-type *Pfkelch13* at full-text screening. Data is extracted into a REDCap [35] database using controlled terminology. Variables assessed for data extraction are listed in **Supplementary Table S2** and **Supplementary Materials S3**.

#### Study eligibility criteria for full-text screening

The following criteria are applied during full-text screening to evaluate a study’s eligibility for inclusion in this proposed IPD meta-analysis.

#### Inclusion criteria

Prospective clinical trials in patients with uncomplicated falciparum malaria should involve at least one study arm receiving treatment with an ACT or artemisinin monotherapy, with the endpoint being the clinical and parasitological response. This response is assessed through:

□ Repeated measures of parasitaemia at least twice daily during the first three days of treatment or until a repeated negative count is obtained, facilitating the calculation of parasite clearance half-life (PC½).
□ Parasite positivity on Day-2 and Day-3.
□ PCR-corrected clinical and parasitological response by Day-28 or later.

Additionally, trials should report the *Pfkelch13* genotype at baseline (before treatment or by Day-3) and ensure the availability of the antimalarial dosing schedule, at least as per protocol.

#### Exclusion criteria

□ Severe malaria on enrolment (participant level) or exclusively (study level).
□ Only reporting on wildtype *Pfkelch13*.
□ Studies that involve induced malaria (healthy volunteer infection studies).
□ Studies that involve malaria in animals.
□ Studies evaluating treatment, such as mass drug administration, healthy volunteers, or prophylaxis or chemoprevention studies.
□ Any study where no non-synonymous *Pfkelch13* mutations are detected.

#### Desirable variables

The following variables are listed as desirable:

□ Molecular markers of partner drug resistance
□ Polymerase Chain Reaction (PCR) genotyping methods to distinguish reinfection and recrudescence.
□ Haemoglobin (Hb) concentration at enrolment.
□ Fever (or body temperature) at enrolment.
□ Gametocyte density at enrolment and any follow-up measurement(s).
□ Patient height.
□ Any comorbidities.
□ Any concurrent medication (e.g. rifampicin).

#### Study metadata

- Study location/s, study year/s, study population/s.
- Transmission intensity (low/moderate/ high) of each study site.
- Study design (e.g. randomised controlled trial, observational study).
- Artemisinin-based treatment/s and dosing strategies (age-based or weight-based; fat coadministration; supervised/partially supervised/unsupervised).
- Microscopy quantification methodology for asexual (and sexual) parasites
- *Pfkelch13* genotyping methods
- Methods for PCR genotyping to distinguish reinfection and recrudescence.

### Data acquisition and data management

#### Collating IPD

Following identifying eligible studies, principal investigators will be contacted to participate in the study group. Researchers agreeing to the submission terms and conditions will be requested to upload de-identified IPD to the WorldWide Antimalarial Resistance Network (WWARN) repository through a secure web portal. For studies already shared with WWARN, permission to include data in the current study will be requested from the principal investigators or the independent Infectious Diseases Data Observatory (IDDO) Data Access Committee [18, 36]. All data are treated in compliance with the UK Data Protection Act (2018, the UK General Data Protection Regulation [UK GDPR] and the European Union General Data Protection Regulation [EU GDPR]).

#### Data integrity, study group governance and ethics

This study utilises the Infectious Diseases Data Observatory (IDDO) data platform, which includes datasets from previously submitted and newly submitted studies uploaded to the platform are owned by the original controllers responsible for ensuring that data was collected per the applicable laws and ethical approval for the countries where the study was conducted. In addition to the de-identification processes completed by data contributors, IDDO performs additional checks before or during data curation to ensure that data are pseudonymised. Before releasing it to data requestors, this process ensures full compliance with international regulations such as the UK Data Protection Regulation (UK GDPR), the European Union Data Protection Regulation (EU GDPR), and the Data Protection Act 2018. More information about IDDO’s data governance can be found on IDDO’s website (iddo.org).”

#### Data contributor’s participation

Data contributors of eligible studies will be invited to become members of the study group and given the opportunity to participate actively in the analysis, interpretation of results, and manuscript preparation. Co-authorship for any publications resulting from these analyses will be assigned per the IDDO publication policy and available via https://www.iddo.org/sites/default/files/2024-11/iddo_publication_policy_final5.pdf [37].

### Statistical Analysis Plan (SAP)

#### Study population characteristics

From the IPD shared, baseline characteristics of patients will be summarised, including those samples with *Pfkelch13* markers (**Table 1**) or samples that are wildtype, by region and transmission intensity and with information on pre-defined covariates including age, sex, nutritional status, baseline parasitaemia, fever, haemoglobin (g/dL) or haematocrit (%), anaemia and severe anaemia and treatment information: artemisinin-based treatment regimen, total mg/kg dose for each artemisinin derivative, dosing strategies, and treatment supervision.

A summary of the characteristics of the identified eligible studies will be compiled, which provides details on:

□ The study location, duration, population, treatment groups, methodologies for parasite quantification, genotyping techniques, and the number of isolates genotyped (**Supplementary Table S2 and S3**).
□ To highlight potential bias, a summary of the eligible trials uploaded to the WWARN repository and eligible studies not shared with WWARN will be presented.
□ A table of the methodology used will be presented. This will include study design, parasitaemia sampling schedules, molecular analysis for *Pfkelch13* markers and PCR correction, artemisinin-based treatment regimen, supervision/food intake with ACT, follow-up duration, study populations, location by country and malaria transmission intensity.

#### Types of interventions, exposure and controls

This IPD-MA exclusively examines studies in which patients with uncomplicated falciparum malaria were treated with either an ACT or an artemisinin-based monotherapy. The primary exposure of interest is the presence of any non-synonymous mutations in the *Pfkelch13* gene of *P. falciparum* (exact list prespecified in **Table 1)**. Controls will be malaria infections with a wild-type only (artemisinin-sensitive) *Pfkelch13* genotype (mixed infections with wild-type and mutant parasites will be treated as mutant infections). We will calculate the counts and proportions (with a 95% confidence interval) of the identified *Pfkelch13* mutations for all studies, stratified by study location, region, and the type of ACT used. Patients exhibiting mixed species-specific genotyping results will be described in a descriptive analysis but excluded from further analysis.

#### Outcomes

The primary outcome is:

1. parasite clearance half-life (PC½), estimated using the WWARN parasite clearance estimator, available via https://www.iddo.org/wwarn/parasite-clearance-estimator-pce [38]

Secondary outcomes are:

1. parasite positivity on Day-2 or Day-3 (i.e. 48- and 72-hours post-artemisinin-based treatment initiation);
2. PCR-corrected therapeutic efficacy by Day-28 or later (as defined in the primary study)

#### Statistical assessment of variables

Their mean and standard deviation will describe the distribution of continuous variables if the data are normally distributed, geometric mean and 95% reference range if the data are normally distributed following a log transformation (Shapiro-Wilk test), or the median and interquartile range [8] if the data are non-normally distributed. In addition, graphical tools such as histograms and QQ plots will be used to assess distributional assumptions. Counts, percentages, and frequency distributions will be provided for categorical variables. Tests of statistical significance will not be undertaken for baseline characteristics; instead, the clinical importance of any differences in the baseline distributions will be noted.

#### Analysis of the primary endpoint: Parasite clearance half-life

The parasite clearance rate will be estimated for each individual using use the WWARN parasite clearance estimator [38] (**Supplementary Material S4**). A table will summarise the distribution of parasite clearance half-life by *Pfkelch13* mutation and stratify it by region, transmission intensity, study site, and artemisinin-based treatment and mutation. This information will be plotted to illustrate the overall distribution of parasite clearance half-life by *Pfkelch13* mutation and stratify it by region and transmission intensity.

The primary analysis will be a one-stage IPD-MA examining the association between all pre-specified mutant genotypes (**Table 1**) and the parasite clearance rate. This will consist of a linear regression model onto clearance rate half-life, adjusted for study site (random effects), participant age, and baseline parasitaemia (fixed effect). As none of the mutant alleles has gone to fixation, we do not expect complete collinearity between alleles and study sites.

### Thresholds for determining phenotypic resistance

Previous work based on IPD from Southeast Asia suggested that a threshold parasite clearance half-life of 5 hours could discriminate between mutant and wild-type infections [48]. This threshold will likely not be generalized to sub-Saharan Africa, where the majority of patients have faster baseline clearance due to differences in immunity. We will estimate the distribution of parasite clearance half-lives using a normal mixture model (as done in White *et al.* [48]) to determine a more appropriate threshold for calling phenotypic resistance. The characteristics of this threshold will be determined on held-out data using 10-fold cross-validation (data partitions will be based on the study site).

#### Analysis of Secondary endpoints

##### Parasite positivity on Day-2 or Day-3

The Proportion Parasite Positive (PPP) is defined as the proportion (%) of patients with asexual parasites detectable by microscopy on Day 2 (and on Day-3). The relationship between *Pfkelch13* markers and positivity on Day-2/3 will be explored using a logistic regression model with the study site as a random effect and the baseline parasitaemia and age as a fixed effect. Additionally, the intra-class correlation in the logistic regression model will be investigated.

##### Risk of Recrudescence

Recurrent infection is defined as the reappearance of the parasite from Day-4, from the start of treatment and after a negative parasite count is recorded until the censored event or the end of follow-up. The proportion of patients with different outcomes will be presented. Recurrent infections will then be classified as recrudescence or reinfection using PCR correction (as per the original study). The risk of recrudescence of *P. falciparum* infection will be summarised using the Kaplan-Meier method, and comparison between groups will utilise a log-rank test (stratified test to take site-specific differences). The relationship of *Pfkelch13* markers with recrudescence of *P. falciparum* infection during the follow-up will be assessed through the Cox proportional hazards regression. Random effects in the form of shared *gamma* frailty for study sites will be used to account for unobserved statistical heterogeneity.

The statistical analyses listed in this protocol will be performed using the R Studio Core Team (2022) (R Foundation for Statistical Computing, Vienna, Austria).

The statistical analysis plan may require further development to accommodate evolving requirements. Depending on data availability, amendments and additional analyses may be necessary to address these emerging needs. An updated SAP will be available online under <u>WWARN Study Groups</u>.

#### Risk of bias assessment

The risk of bias judgments for each study will utilize the Cochrane Risk of Bias tool (version 2.0 [39]) for randomized studies and the ROBINS-I tool [40] for non-randomized studies. These judgments will be integrated into our analyses by performing subgroup analyses among studies and participants with a low overall risk of bias or through formal interaction analyses based on the item-level risk of bias responses.

The overall certainty of evidence will be judged based on GRADE guidelines that offer explicit criteria for evaluating the quality of evidence, including the risk of bias, imprecision, inconsistency, indirectness, and publication bias [41].

### Ethics and dissemination

Data have been obtained with informed consent and ethical approvals applicable to the countries where studies were conducted and pseudonymised before or during curation within the IDDO/WWARN repository. While data ownership remains with the data contributor, researchers can request access to data in the WWARN repository. This IPD meta-analysis will address scientific questions similar to the original research questions. This IPD meta-analysis met the criteria for waiver of ethical review as defined by the Oxford Tropical Research Ethics Committee (OxTREC), as the research consists of a secondary analysis of existing pseudonymised data shared in a controlled environment [52].

Findings will be reported following the PRISMA-IPD guidelines. Any publications based on the findings of this IPD meta-analysis will be following the guidelines of the International Committee of Medical Journal Editors [34, 42].

## Discussion

This study protocol builds on the foundational work established by the WWARN *Pfkelch13* Genotype-Phenotype Study Group [8]. Here, we detail the pragmatic search strategy and robust SAP in this protocol, which is the first of its kind. It is geared to assess IPD using a meta-analysis specifically curated to explore the prevalence, distribution, and functional and clinical significance of *Pfkelch13* markers in *P. falciparum* populations globally. The protocol and analyses are ideal for evidence synthesis as they allow exploration of different risk factors, otherwise impossible through an aggregate data meta-analysis alone [42]. As the global ART-R landscape continues to evolve, this protocol aims to serve as a model for further standardised statistical methodologies and definitions for subsequent IPD meta-analyses, promoting its reproducibility in future studies and potentially regular updates.

This protocol responds to the dire need for an updated investigation to include African-specific data, as minimal data was available before ART-R emergence in Africa and thus not included in previous IPD meta-analyses [8, 11]. However, given that ART-R has recently emerged in Africa, and the region accounts for 95% of the global malaria burden [2, 43], an improved understanding of the genetic determinants driving ART-R and its clinical consequences is of paramount importance. The recent and independent emergence of *ART-R* genotypes in East Africa was observed in parallel with diminished ACT efficacy [4, 5, 15, 44–50]. Such regional insights are imperative as the current global genomic surveillance greatly relies on data from South-East Asia [8, 11] [9], from which the WHO classification as “validated” or “candidate” genetic markers was derived [9] and inadequately represents the genetic diversity of African *Plasmodium* populations.

Applying the outlined protocol will generate up-to-date data to help strengthen and enhance the efficiency of ART-R surveillance and response. By slowing parasite clearance, *Pfkelch13* may facilitate reduced ACT efficacy; however, ART-R also depends on the partner drug’s effectiveness [50]. Insight into the association between specific *Pfkelch13* mutations is crucial, as when ART-R emerged previously in the Southeast Asia region, it soon became coupled with resistance to ACT partner drugs, resulting in ACT treatment failures [51–56]. A similar trajectory in sub-Saharan Africa could jeopardize the effective treatment of hundreds of millions of malaria patients annually. A repeat of the increase in malaria morbidity and mortality, seen historically with chloroquine resistance [28], can only be avoided by promptly detecting up-to-date ART-R markers and coordinating efforts to mitigate their impact.

In conclusion, this protocol provides a detailed plan for analysis of the global distribution of *Pfkelch13* markers and their impact on malaria treatment outcomes. It also helps inform an updated categorization of *Pfkelch13* markers of ART-R. This work is critical for refining malaria molecular marker surveillance and optimizing malaria treatment strategies globally, particularly in sub-Saharan Africa, where the burden of malaria remains disproportionately high.

## Data Availability

All data produced in the present study are available upon reasonable request to the authors

## Declarations

### Author contributions

JCV, SvW, PD, KB and PJG designed the systematic review and IPD meta-analysis. SvW, JCV, and SA literature search the studies. SvW and KB wrote the first draft. SvW, SAD, KB, PD, PJG, and JAW contributed to the manuscript.

## Acknowledgements

We appreciate the advice of Joanna Czekajska and Caitlin Richmond. Many thanks to Eli Harriss and Farhad Shokraneh for performing the literature search. Also, thanks to Abdalla Munir and other colleagues at IDDO, who extracted the data in the first version of this study and to members of the WorldWide Antimalarial Resistance Network (WWARN) K13 Study Group.

## Funding

The Global Health EDCTP3 Joint Undertaking supports the MARC SE-Africa, and its members are established under the European Union’s research and innovation programme, Horizon Europe (grant number and project number: 101103076). Grants from the ExxonMobil Foundation and the Bill & Melinda Gates Foundation funded this pooled analysis. Funders had not provided input on the study’s planning, analysis, or presentation.

Disclaimer: Views and opinions expressed are, however, those of the authors only and do not reflect those of the EU or the GH EDTP3 JU. Neither the EU nor GH EDCTP JU can be held responsible for them.

## List of abbreviations

ACT: Artemisinin-based Combination Therapies
AL: Artemether-Lumefantrine
ART-R: Artemisinin resistance
CDISC: Clinical Data Interchange Standards Consortium
DRC: Democratic Republic of the Congo
EU GDPR: European Union General Data Protection Regulation
GRADE: Grading of Recommendations Assessment, Development and Evaluation
Hb: Haemoglobin
HL: Parasite clearance half-life*
IDDO: Infectious Diseases Data Observatory
IPD: Individual Patient Data
IPD-MA: Individual Patient Data Meta-Analyses
MARC SE-Africa: Mitigating Antimalarial Resistance Consortium in South-East Africa
PC½: Parasite Clearance Half-life
PCR: Polymerase Chain Reaction
*PfKelch13*: *Plasmodium falciparum Kelch* 13
PPP: Proportion Parasite Positivity
PRISMA-IPD: Preferred Reporting Items for Systematic Review and Meta-Analyses of individual participant data
ROC: Receiver Operating Characteristic
RSA: Ring-stage Survival Assay
SAP: Statistical Analysis Plan
TES: Therapeutic Efficacy Studies
UK: United Kingdom
WCTL: WWARN Clinical Trial Library
WHO: World Health Organization
WWARN: WorldWide Antimalarial Resistance Network
WWARN DMSAP: WWARN Clinical Module Data Management and Statistical Analytical Plan
WWARN PCE: WWARN Parasite Clearance Estimator

*HL is used instead of PC½ for statistical analyses to remain consistent and align with the terminology of published statistical literature.

