## Supplemental_materials for "A protocol for a systematic review and individual patient data meta-analysis investigating the relationship between *Pfkelch13* mutations and response to artemisinin-based treatment for uncomplicated falciparum malaria": Supplementary_Materials_SvW_PD.docx

**Supplementary Material S1:**

**Standardised Protocol Definitions**

**Antimalarial dose:** The doses of artemisinin derivatives and partner compounds received will be calculated from the daily tablets administered to each patient. If the daily tablet counts are unavailable, doses will be estimated retrospectively using the dosing scheme provided in the study protocols.

**Malaria transmission intensity:** Study sites will be categorized into three groups: low, medium, and high malaria transmission, according to parasite prevalence estimates provided by the Malaria Atlas Project, tailored to the specific locations and years when participants were enrolled [1].

**Anaemia:** Anaemia is a haemoglobin (Hb) level below 10 g/dL. Severe anaemia is characterized by a Hb level of less than 7 g/dL.

**Parasite clearance half-life:** Parasite clearance half-life is the time required for parasitaemia to be reduced by half (50%) during the log-linear phase of parasite clearance [2, 3]. The PC½ is closely connected to the parasite clearance rate, which is defined as the negative of the slope of the log-parasitaemia profile over the time in which the anti-malarial is having its primary effect [3-5]. Parasite Half-life or HL for statistical analyses is used instead of PC½ to remain consistent and align with the terminology of published statistical literature.

**Fever:** The presence of fever is defined as body temperature > 37.5°C.

**Treatment supervision:** Schizontocidal treatment will be classified as supervised if all doses were directly observed, partially supervised if at least the morning doses of a twice-daily regimen were observed and not supervised if fewer or no doses were observed.

**Recrudescence:** Recrudescence of parasitaemia by the end of the study follow-up is defined as recurrent infections caused by the initial parasites, as confirmed by PCR genotyping. Recrudescence will be differentiated from new infections according to the methods used in the primary study.

***PfKelch13* genotype**: Genotyping results will be based on methods used in the primary study to amplify the *Kelch13* gene region’s propeller domain (codon 400 to 727 on chromosome 13 of 3D7 isolate [PF3D7_1343700]), which is determined via DNA sequencing, as described previously [3].

**Synonymous mutations:** a DNA mutation occurring in the *Pfkelch13* gene that alters the DNA sequence. However, the DNA change does not result in changes in the protein's amino acid sequence once translated. These mutations may not be functionally consequential and are neutral, having no direct effect on the protein's function.

**Non-synonymous mutations**: mutations that result in changes in the DNA sequence that lead to an amino acid substitution in the *Pfkelch13* protein. These mutations can alter the structure or function of the protein, potentially contributing to changes in the parasite’s resistance to artemisinin-based treatments.

**Supplementary Table S1**: Sources for data acquisition and literature searches.

| **Bibliographic Databases** | **Clinical and Research Data Repositories** |
| --- | --- |
| MEDLINE | ClinicalTrials.gov  Pan-African Clinical Trials Registry |
| Embase | WWARN Molecular Surveyor |
| Global Health | WWARN Molecular Database |
| Scopus | WWARN Clinical Trial Libraries |
| Web of Science |  |
| Cochrane Database of Systematic Reviews |  |
| PubMed |  |

**Table S2:** Covariates to be considered for inclusion by outcome

|  | **Parasite clearance half-life (PC1/2)** | **Parasite positivity on Day-2/3** | **PCR corrected / uncorrected ACPR rate** |
| --- | --- | --- | --- |
| **Fixed effects** | Age | Age | Age |
|  | Sex | Sex | Sex |
|  | Baseline parasitaemia | Baseline parasitaemia | Baseline parasitaemia |
|  | ACT | ACT | ACT |
|  | Artemisinin dose^1^ | Artemisinin dose | Artemisinin dose |
|  | Treatment supervision^2^ | Treatment supervision^2^ | Treatment supervision^2^ |
|  | Post-dose vomiting | Post-dose vomiting | Post-dose vomiting |
|  | Nutrition status^3^ | Nutrition status | Nutrition status |
|  | Transmission intensity^4^ | Transmission intensity | Transmission intensity |
|  |  |  | Baseline gametocytaemia |
|  |  |  | Baseline fever |
|  |  |  | Fat administration with ACT |
|  |  |  | Molecular marker/s of partner drug resistance |
|  |  |  | Duration of follow-up |
|  |  |  | PCR correction method |
| Random effects | Study-site | Study-site | Study-site |

1. Actual mg/kg dose, if available, otherwise as per protocol
2. Supervised/partially supervised/unsupervised
3. For children: WHZ score or MUAC if available, otherwise WAZ score
4. Transmission Intensity of the study sites will be classified into 3 categories: low, medium and high malaria transmission based on the parasite prevalence estimates obtained from the [Malaria Atlas Project](https://data.malariaatlas.org/maps?layers=Malaria:202406_Global_Pf_Parasite_Rate) for specific location and year of study.

**Supplementary material S2**: Search terms per repositories, databases, and libraries

### **Information sources and search strategies**

##### WCTL

Below is additional information on the WorldWide Antimalarial Research Network (WWARN) Clinical Trial Publication Library (WCTL; available at www.iddo.org/wwarn/wwarn-clinical-trials-publication-library). This library serves as an important source of curated literature on TES outcomes and *PfKelch* 13 genotyping results. Moreover, it constitutes a comprehensive compilation of all antimalarial clinical efficacy trials conducted and published since 1946 and is updated biannually. It was used for literature searches and systematic literature reviews.

- The following databases are searched for WCTL:

MEDLINE, EMBASE, Web of Science (all Databases), Cochrane Central, WHO Global Index Medicus and Clinicaltrials.gov.

No restrictions are placed on language or publication date. Briefly, the search terms used in the strategy included “Malaria,” “malaria.ti,ab.”, “*Plasmodium*,” “plasmodium.ti,ab.”, “falciparum,” the names of each component of the antimalarial drug, and other related terms. A librarian provided a list of studies identified through the search results provided by the Bodleian Libraries librarian. These were uploaded into Covidence screening software and independently screened by two reviewers using the agreed inclusion and exclusion criteria. The final list of eligible studies is agreed upon, and two data extractors extract the study data into a REDCap database. The extracted data is available via the WWARN website to the broader malaria community.

Our librarian performs literature searches through the Bodleian Library at Oxford University. The library's last update was on 31/08/2024.

**WCTL literature search terms:**

| **Disease & Species Terms Included:** | | | | | | | | |
| --- | --- | --- | --- | --- | --- | --- | --- | --- |
| malaria | *falciparum* | | *vivax* | | *ovale* | | | *knowlesi* |
| *malariae* | *plasmodium* | |  | |  | | |  |
| **Drug Terms Included:** | | | | | | | | |
| ACT-451840 | | Amodiaquine | | Amopyroquin | | AQ-13 | Arteether | |
| Artefenomel | | Arteflene | | Artemether | | Artemether-Lumefantrine | Artemether-Lumefantrine-Amodiaquine | |
| Artemisinin | | Artemisinin-naphthoquine | | Arterolane | | Arterolane-piperaquine | Artesunate | |
| Artesunate-Amodiaquine | | Artesunate-Amodiaquine-Chlorpheniramine | | Artesunate-Mefloquine | | Artesunate-Piperaquine | Atoguanil | |
| Atovaquone | | Atovaquone-Proguanil | | Azithromycin | | Berberine | Bulaquine | |
| CDRI 97/78 | | Chloroquine | | Chlorpheniramine | | Chlorproguanil-dapsone | Chlorproguanil | |
| Cipargamin | | Clindamycin | | Cotrimoxazole | | Dapsone | Dihydroartemisinin- Piperaquine | |
| Dihydroartemisinin- Piperaquine-Trimethoprim | | Dihydroartemisinin | | Doxycycline | | Elubaquine | Erythromycin | |
| Ferroquine | | Fosmidomycin | | Fosmidomycin-piperaquine | | Ganaplacide | Ganaplacide-Lumefantrine | |
| GSK701 | | Halofantrine | | Imatinib | | INE963 | Ketotifen | |
| L9LS antibody | | Lotilaner | | Lumefantrine | | M5717 | M5717-pyronaridine | |
| MMV533 | | Mefloquine | | Metakelfin | | Methylene Blue | Methylene Blue-Amodiaquine | |
| Naphthoquine | | Norfloxacin | | P218 | | Pafuramidine | Pentaquine | |
| Piperaquine | | Primaquine | | Probenecid | | Proguanil | Pyrimethamine | |
| Pyronaridine | | Quinacrine | | Quinidine | | Quinine | Rifampicin-Cotrimoxazole-Isoniazid | |
| Rosiglitazone | | Ruxolitinib | | Sevuparin | | SJ733 | Spiramycin | |
| Sulfadoxine | | Sulfadoxine-Pyrimethamine | | Sulfadoxine-Pyrimethamine-Amodiaquine | | Sulfamethoxazole | Sulfamonomethoxine | |
| Sulfamethoxypyrazine | | Tafenoquine | | Tetracycline | | Trimethoprim | XTB-31F mAB | |
| ZY-19489 | | ZY-19489-ferroquine | |  | |  |  | |

The WCTPL includes randomised control trials, quasi-randomised controlled trials, case-control studies, and longitudinal cohort studies. Pharmacokinetic studies using drugs that are components of artemisinin-based combination therapy are also included. Animal studies, prevention studies, case reports, case series, systematic reviews, and literature reviews were excluded.

WCTL includes studies with at least 28 days of follow-up. Since the primary focus of this meta-analysis is parasite clearance evaluated in the first few days after starting antimalarial treatment, both studies within WCTPL and studies screened but excluded from WTPL were considered for inclusion. Two independent reviewers (SvW and SAD) screened the title, abstract, and full text as necessary, and a third reviewer (EM) was used to resolve any discordances.

#### **Scoping Review search terms**

For the updated review we included a scoping search was performed using PubMed with the following search terms: (PUBMED) (("K13"[All Fields] OR "Kelch 13"[All Fields] OR "Pfk13"[All Fields] OR "kelch13"[All Fields] OR "Pfkelch13"[All Fields]) AND "resistant*"[All Fields] AND ("malaria*"[All Fields] OR "antimalaria*"[All Fields])) AND ({date} [pdat]) AND ("artesunate*"[All Fields] OR "artesunate SP*"[All Fields]))

#### **Clinical trial repository literature search terms**

**Clinicaltrials.gov and The Pan African Clinical Trials Registry (PACTR)**

Search terms listed above were applied to the clinical trials repositories. The exclusion criteria included the following: NOT Prevention NOT chemoprevention NOT pregnancy NOT LLIN NOT insecticide NOT vector control NOT Nets NOT screens NOT Mass Drug Administration NOT vector NOT vaccine NOT RTS,S NOT R21

**Supplementary Materials S3**: Variables assessed for data extraction

| **Register** |  |
| --- | --- |
| **Publication detail** |  |
| Accession number |  |
| Authors of the Publication |  |
| Author-Year (e.g. Rahmasari-2023) |  |
| Year of publication |  |
| Title of publication |  |
| Journal of publication |  |
| Journal Volume |  |
| Issue Number |  |
| Pages of Article |  |
| Date of extraction |  |
| Clinical Trials Registration Number |  |
| Is this a clinical trial registration without published results? | Yes |
|  | No |
| Reported Deaths in the Study | Yes |
|  | No |
|  | Unspecified |
| Exclusion/Inclusion Reason |  |
| Comments |  |
| **Location information** |  |
| Country |  |
| Study site |  |
| Exclusion/Inclusion Reason |  |
| **Study Data** |  |
| First Year of Patient Recruitment |  |
| Last Year of Patient Recruitment |  |
| Study type: | INTERVENTIONAL |
|  | OBSERVATIONAL |
| What is the study type (please select one) | ADHESION PERFORMANCE |
|  | ALCOHOL EFFECT |
|  | BIO-AVAILABILITY |
|  | BIO-EQUIVALENCE |
|  | BIOSIMILARITY |
|  | DEVICE-DRUG INTERACTION |
|  | DIAGNOSIS |
|  | DOSE FINDING |
|  | DOSE PROPORTIONALITY |
|  | DOSE RESPONSE |
|  | DRUG-DRUG INTERACTION |
|  | ECG |
|  | EFFICACY |
|  | FOOD EFFECT |
|  | IMMUNOGENICITY |
| What is the purpose of the study (as reported in the publication)? | IMMUNOGENICITY |
|  | PHARMACODYNAMIC |
|  | PHARMACOECONOMIC |
|  | PHARMACOGENETIC |
|  | PHARMACOGENOMIC |
|  | PHARMACOKINETIC |
|  | POSITION EFFECT |
|  | PREVENTION |
|  | REACTOGENICITY |
|  | SAFETY |
|  | SWALLOWING FUNCTION |
|  | THOROUGH QT |
|  | TOLERABILITY |
|  | TREATMENT |
|  | USABILITY TESTING |
|  | WATER EFFECT |
| Was it an intervention compared to another intervention? | YES |
|  | NO |
| Was the study randomized? | YES |
|  | NO |
|  | YES |
| Was the study blinded? | NO |
| What was the observational model used? | CASE CONTROL |
|  | CASE CROSSOVER |
|  | CASE ONLY |
|  | COHORT |
|  | ECOLOGIC OR COMMUNITY |
|  | FAMILY BASED |
| What was the observational model time perspective used (Select one)? | CROSS SECTIONAL |
|  | PROSPECTIVE |
|  | RETROSPECTIVE |
| Was there a phase for the clinical trial? If so please state | UNKNOWN |
|  | PHASE 0 TRIAL |
|  | PHASE I TRIAL |
|  | PHASE I/II TRIAL |
|  | PHASE II TRIAL |
|  | PHASE II/III TRIAL |
|  | PHASE IIA TRIAL |
|  | PHASE IIB TRIAL |
|  | PHASE III TRIAL |
|  | PHASE IIIA TRIAL |
|  | PHASE IIIB TRIAL |
|  | PHASE IV TRIAL |
|  | PHASE V TRIAL |
| What were the stated inclusion criteria? |  |
| What were the stated exclusion criteria? |  |
| **Study Data Participant Information** |  |
| What was the planned minimum age of participants? |  |
| What was the planned minimum age of participants? | DAYS |
|  | MONTHS |
|  | YEARS |
| What was the planned maximum age of participants? |  |
| What was the planned maximum age of participants - unit? | DAYS |
|  | MONTHS |
|  | YEARS |
| Did the study recruit subjects younger than 5 years of age? |  |
| Did the study recruit subjects ages 5 to 15 years? |  |
| Did the study recruit subjects older than 15 years? |  |
| Did the study recruit Pregnant Women? |  |
| Length of Follow-up (in Days) |  |
| *Plasmodium falciparum included in the study?* |  |
| Mixed Infection (Pf+Pv) included in the study? |  |
| Species besides Pf? Please specify |  |
| Does this study contain an ACT? Please specify |  |
| **Treatment** |  |
| How many treatment arms are there in this study? |  |
| **Please complete this for each treatment arm by copying and pasting "Treatment" queries (Rows 115-141) in subsequent columns until all treatment arms are captured** | |
| Number of Patients Recruited in this Treatment Arm |  |
| Is the number of patients recruited in this arm the intention-to-treat population or the per-protocol population? | |
| Verified Treatment Name |  |
| Was the drug administration supervised in this Treatment Arm? | Fully Supervised |
|  | Unsupervised |
|  | Partially supervised |
|  | Not Specified |
| Formulation of Drugs in this Treatment Arm |  |
|  | Fixed-Dose Combination |
|  | Loose |
|  | Co-blister Pack |
|  | Not Specified |
| Manufacturer of Drugs in the Treatment Arm |  |
| Is information on the drug batch number or lot number available for drugs in this Treatment Arm? |  |
| Name of Drug 2 in this Treatment Arm |  |
| Name of Drug 3 in this Treatment Arm |  |
| Is there an ACT in this Treatment Arm? |  |
| **Species Assessed at this Site** |  |
| Site Information |  |
| Country |  |
| Location of the study |  |
| Site Coordinates |  |
| Latitude of this location |  |
| Longitude of this location |  |
| Endemicity of transmission area |  |
| **Molecular and Phenotypic variables** |  |
| **Molecular Markers** |  |
| What molecular markers were used for genotyping? | MSP1, MSP2, and microsatellites |
|  | MSP1, MSP2, and glurp |
|  | SNP barcodes |
|  | Amplicon sequencing |
|  | Other (specify) |
| List all K13 genotypes investigated (e.g. R561H) |  |
| List all K13 genotypes present/ prevalent (e.g. R561H) |  |
| **Molecular Correction Method** |  |
| What method was used to distinguish new infection from recrudescence (Molecular correction method)? | 3/3 Approach (WHO / MMV) |
|  | 2/3 Approach |
|  | Bayesian analysis |
|  | Other (specify) |
| Statistical Analysis Method |  |
|  | What statistical analysis method was used to calculate PCR-corrected efficacy? |
|  | Kaplan-Meier survival analysis |
|  | Per protocol analysis |
|  | Other (specify) |
| **Phenotypic Variable** |  |
| Was a phenotypic variable observed (Please select) | Delayed parasite clearance |
|  | TES outcomes |
|  | Parasite clearance half-life (PC1/2) in patients with falciparum malaria treated with ACTs or monotherapy in antimalarial clinical studies by location, treatment, study population and date. |
|  | Parasite positivity on Day 2 and Day 3 in patients with falciparum malaria treated with ACTs or monotherapy in antimalarial clinical studies by location, treatment, study population and date. |
|  | Clinical and parasitological treatment response in patients with falciparum malaria treated with ACTs or monotherapy in antimalarial clinical studies by location, treatment, study population and date. |

**Supplementary Materials S4**: **Statistical Analyses**

*Regression modelling for identifying mutations associated with parasite clearance half-life*

The analyses of HL will be conducted using a one-stage IPD-MA: The one-stage IPD-MA will be undertaken separately by region and transmission intensity, respectively. In the IPD-MA, individual patient HL estimated from the WWARN PCE tool will be modelled after log transformation. Linear regression models will be fitted, with *Pfkelch13* markers, age, treatment, total mg/kg artesunate dose, fever, initial parasitaemia, and presence of gametocytes on enrolment as covariates. Random effects for the study site will be used to account for heterogeneity between studies. Appropriate diagnostics, such as residuals, will be examined to assess the underlying model assumption, such as normality, and to identify any systematic departure from the model assumptions.

Wald test will be used to assess the difference between individual *PfKelch13* markers and the wild type, and the percentage difference in HL (95% CI) will be calculated as an exponent of the difference of the corresponding regression coefficients. For the univariate model, specific outputs to be presented for the covariates (*PfKelch13* markers) included in the model include regression coefficients and associated 95% CI. The covariates significantly (statistically) impacting the log HL will not contain zero in their 95% CI. The significance level of α = 0.05 will be devised by the number of covariates in the case to manage multiple comparisons through Bonferroni-corrections.

*Determination of appropriate HL threshold*

A finite mixture model will be fitted with two components (sensitive and resistant) to characterise parasite clearance distribution in the populations included. Further explorations will be examined, including the utility of the Receiver Operating Characteristic (ROC) curve to define an HL cut-off. The patients with infections and *PfKelch13* markers associated with an increase in HL (compared with the wild-type group, adjusted for covariates) will form group 1, and all other patients will be in group 0 for the evaluation of resistance. Thus, the ROC curve will be plotted, and the area under the ROC curve will be calculated with its standard error as described previously [7] and its exact binomial confidence interval. From this HL, a cut-off of two population groups will be described to differentiate slow-clearing populations, defined as all isolates with markers associated with a significant increase in HL values in this analysis. In contrast, the fast-clearing population will include all other isolates. Their correspondence to the parasite positivity proportion on Day-3 and *PfKelch13* markers will be investigated.

**Supplementary Material S5:** **Covariate selection and regression models**

Multivariable regression modelling will be carried out as outlined previously using a general approach recommended in the literature [8]. Briefly, the variable selection approach will consider using a likelihood ratio test (a backward elimination approach is preferred) [8]. To find the most plausible model, models will be compared using information criteria such as the Likelihood Ratio Test (LRT) for nested models and Akaike’s Information Criterion (AIC) for non-nested models.

*Missing data*

The regression analysis will exclude variables with more than 50% missing data. The underlying distribution of missingness will be investigated to gauge potential missingness mechanisms and guide appropriate analysis models. For example, under covariate-dependent missingness (missing at random assumption), multiple imputation will be considered [9].
